# Optimal disease surveillance with graph-based Active Learning

**DOI:** 10.1101/2024.06.21.24309284

**Authors:** Joseph L.-H. Tsui, Mengyan Zhang, Prathyush Sambaturu, Simon Busch-Moreno, Marc A. Suchard, Oliver G. Pybus, Seth Flaxman, Elizaveta Semenova, Moritz U. G. Kraemer

**Affiliations:** Department of Biology, University of Oxford, UK; Department of Computer Science, University of Oxford, UK; Department of Biostatistics, University of California, Los Angeles, CA, US; Department of Pathobiology and Population Science, Royal Veterinary College, UK; Pandemic Sciences Institute, University of Oxford, UK; Department of Epidemiology and Biostatistics, Imperial College London, UK

## Abstract

Tracking the spread of emerging pathogens is critical to the design of timely and effective public health responses. Policymakers face the challenge of allocating finite resources for testing and surveillance across locations, with the goal of maximising the information obtained about the underlying trends in prevalence and incidence. We model this decision-making process as an iterative node classification problem on an undirected and unweighted graph, in which nodes represent locations and edges represent movement of infectious agents among them. To begin, a single node is randomly selected for testing and determined to be either infected or uninfected. Test feedback is then used to update estimates of the probability of unobserved nodes being infected and to inform the selection of nodes for testing at the next iterations, until a certain resource budget is exhausted. Using this framework we evaluate and compare the performance of previously developed Active Learning policies, including node-entropy and Bayesian Active Learning by Disagreement. We explore the performance of these policies under different outbreak scenarios using simulated outbreaks on both synthetic and empirical networks. Further, we propose a novel policy that considers the distance-weighted average entropy of infection predictions among the neighbours of each candidate node. Our proposed policy outperforms existing ones in most outbreak scenarios, leading to a reduction in the number of tests required to achieve a certain predictive accuracy. Our findings could inform the design of cost-effective surveillance strategies for emerging and endemic pathogens, and reduce the uncertainties associated with early risk assessments in resource-constrained situations.

## Introduction

Infectious disease surveillance is necessary for managing infectious disease outbreaks, enabling public health authorities to monitor and respond to ongoing disease spread. Notable examples in the past decade include the 2014-2016 West African and 2018-2020 Kivu Ebola virus epidemics, and the COVID-19 pandemic, for which the early detection and continued tracking of the virus’ spread helped to inform the design of interventions including targeted vaccination (1–5), case isolation (6–10) and social distancing (11–14). Without timely and accurate surveillance data, the effectiveness of these interventions would likely have been compromised. For example, travel restrictions targeted at countries where new variants of SARS-CoV-2 were first observed were rendered largely ineffective by delay in case detection and insufficient pathogen sequencing (15, 16). Similarly, the lack of baseline testing prior to the 2015-2016 Zika virus epidemic in the Americas likely contributed to the delay in the identification of the scale of disease spread, thereby allowing the virus to disseminate to new locations before a coordinated response was initiated (17, 18).

Well documented examples of effective disease surveillance have been limited largely to within-country initiatives (e.g., the Real-time Assessment of Community Transmission (REACT) in the UK (19), and the National Notifiable Diseases Surveillance System (NNDSS) in the US (20), while globally-coordinated programs remain rare (21). This can lead to a disproportionate or inequitable distribution of testing resources within and between regions or countries, with some locations able to conduct large-scale mass testing for sustained periods of time, while others manage only sparse or sporadic testing (22, 23). One study showed that the intensity of virus genomic sequencing during the COVID-19 pandemic was positively associated with Research and Development expenditures at a country level (24). This likely allowed the virus to continue proliferating undetected in locations with insufficient testing, potentially prolonging local outbreaks.

Previous research on infectious disease surveillance has focused primarily on developing models to identify sentinel sites or subpopulations, with the objective of classifying nodes in networks that could serve as observational units for monitoring disease spread (25–27). Since the COVID-19 pandemic, there has been growing interest in the design of optimal control measures to contain transmission (28), with some studies examining the cost-effectiveness of different strategies for testing and isolation (29–32); one recent study also explored the impact of different air travel regulations on the likelihood of a local epidemic escalating into a global pandemic (33). However, the effectiveness of these interventions depends ultimately on the capacity of local authorities to conduct surveillance and to collectively provide an accurate assessment of overall disease distribution at any stage of an outbreak - a challenge which, to the best of our knowledge, has received little attention to date (34).

This study attempts to address this question - specifically, we consider how testing should be performed across a mobility network, with the objective of providing accurate estimates of where a disease is present, given a fixed budget of testing resources. We hypothesize that the design of an appropriate policy for this task can be formulated as a node classification problem with Active Learning (AL). This motivates the development of an adaptive test deployment framework, which we use to evaluate and compare the performance of previously developed AL policies for infectious disease surveillance. We further propose a novel policy that takes into consideration graph-based uncertainties, named *Selection by Local-Entropy (LE)*, which we show outperforms similar existing policies in most outbreak scenarios and on networks with a diverse range of structural properties, including those commonly found in empirical human mobility networks.

## Materials and Methods

### Disease Surveillance as a Node Classification Task

We consider the deployment of a disease surveillance program on a mobility network as a node classification task, with the mobility network representing the movement patterns of individuals in a population. The goal of a policymaker (or agent) in this classification task is to predict the presence or absence of a disease of interest (or whether disease prevalence is above or below a certain threshold) at any unobserved node, given knowledge of the infection status at a subset of nodes. We represent the mobility network as an undirected and unweighted graph *G* = (*V, E*), with *N* nodes *v_i_* ∈ *V* representing locations, and edges (*v_i_, v_j_*) ∈ *E* representing the existence of movement of infectious agents between nodes *v_i_* and *v_j_*.

Prior to the start of a surveillance program, we assume that there is an underlying distribution of infections resulting from an outbreak that originated from a single infected node. Importantly, we assume that i) the outbreak can be modelled as a stochastic Susceptible-Infected (SI) process, with transmission occurring only between an infected node and an uninfected node if there is an edge between them (see S.1 in Supplementary Information (SI) for further details), and ii) that the timescale over which transmission occurs is sufficiently longer than the timescale over which testing is deployed, such that the underlying disease distribution can be considered to be static over the course of the surveillance program. To indicate the underlying disease distribution, each node *v_i_* in the mobility network is assigned a binary label *y_i_* ∈ {0, 1} representing its infection status, where *y_i_* = 1 if the node is infected (disease presence) and *y_i_* = 0 if uninfected (disease absence). Exploration of the impact of relaxing these assumptions will be addressed in future work.

Here we assume that each surveillance program begins with the known infection status of a single (randomly selected) node, with nodes of either infection status being equally likely to be selected. Given this initial observation, a hypothetical policymaker is then tasked with answering the question: *how should a finite amount of testing resources be deployed across the network to maximise the information gained about the underlying disease distribution?*

### Test Allocation as an Active Learning Task

The study of this question is known as Active Learning (AL) (35), during which the objective is to maximise the predictive performance of a model in training (also known as a surrogate model) with the fewest possible instances of labelled data. The process of labelling data is often expensive and time-consuming, and therefore the selection of data instances for labelling can be alternatively performed in an iterative fashion such that a small subset of unlabelled data instances are selected at each iteration by an AL acquisition policy (referred to as an allocation policy hereafter). The labels of this subset are then revealed and used as input to retrain the surrogate model and to generate label predictions for remaining unlabelled instances. Note that the label of each node is assumed to be unchanging between iterations. Additionally, we assume that a single node is selected at iteration.

There exist a number of previously developed approaches for constructing the surrogate model, e.g., label propagation (36), Gaussian Random Fields (37), and Graph Neural Networks (GNN) (38). Here we adopt an approach that is popular in spatial epidemiology known as the Conditional Autoregressive (CAR) model (39), which assumes that the value of a variable at a given node in a network is conditional on the values at neighbouring nodes, with weights specified by an adjacency matrix, ***A***. In the context of disease surveillance with binary infection status, the CAR model allows us to estimate *p*(*v_i_*|***D_r_***) for a given node *v_i_*, i.e. the probability that the node is infected conditional on the observed data ***D_r_*** = {(*v_j_, y_j_*)|∀*v_j_* ∈ *V_obs,r_*}, where *y_j_* is the observed infection status of a node *v_j_*, and *V_obs,r_* is the set of nodes with known infection status up to the current iteration *r* (see S.2 in SI for a detailed description of the model). Note that it is not the focus of this study to consider the predictive performance of different surrogate models, but rather the relative performance of different test allocation strategies given the same surrogate model. In future extensions where additional model complexities are incorporated (e.g., weighted and directed edges, node attributes), the performance of the different strategies given different surrogate models should be compared and assessed.

At each iteration, the predicted infection status generated by the surrogate model are used to guide the selection process at the next iteration, according to the allocation policy of choice. One important group of AL policies is uncertainty-based, i.e. they select nodes for observation according to where the surrogate model’s predictions are maximally uncertain. One common measure of uncertainty is the information entropy of label predictions: the larger the entropy, the more uncertain the model is about its label prediction for a given node, and the more likely it is that the node would be selected for observation at the next iteration. Another state-of-the-art uncertainty-based policy is Bayesian Active Learning by Disagreement (BALD) (40), which selects nodes that maximise the mutual information between label predictions and model posterior (see S.3 in SI for more details). A number of alternatives to uncertainty-based policies exist in the AL literature, e.g., graph-based heuristics and Expected Error Reduction (41). In this study we focus primarily on policies that rely on graph-based heuristics and uncertainty-based policies that are adaptive (i.e. policies that select nodes iteratively using information from previous observations; see Table 1 for a summary of all policies considered in this study.)

**Table 1:** Summary of policies considered in this study. Abbreviation for each policy is shown in brackets following the policy name. For all policies, random tie-breaking is performed if and when there are multiple candidate nodes given equal preference according to a selection criterion.

| Allocation Policy | Policy Type | Brief Description |
| --- | --- | --- |
| Least-Confidence (LC) (42) | - Uncertainty-based<br>- Adaptive | Select the unlabelled node with predicted infection probability (posterior mean) that is closest to 0.5, indicating the least confidence in label prediction. |
| Node-Entropy (NE) (42) |  | Select the unlabelled node with the highest entropy in its label prediction according to the surrogate model. It can be shown that NE always selects the same node as the policy LC at any iteration (see S.4 in SI); as a result, only NE is considered hereafter. |
| Bayesian Active Learning by Disagreement (BALD) (40) |  | Select the unlabelled node with the highest mutual information between label prediction and posterior from the surrogate model. |
| Local-Entropy (LE)<br>(our proposed policy) | | Select the unlabelled node with the highest *Local-Entropy*, as defined by Equation (1)–(3), with $\lambda = 0$ (maximal exploration). |
| Degree-Centrality (DC) | - Graph-based<br>- Non-Adaptive | Select the unlabelled node with the highest degree-centrality (most connections). |
| PageRank-Centrality (PC) |  | Select the unlabelled node with the highest PageRank-centrality (43). |
| Reactive-Infected (RI) | - Benchmark<br>- Adaptive | Select at random an unlabelled node among immediate neighbours of nodes that are known to be infected from previous observations, if available; otherwise, sample randomly from remaining unlabelled nodes. |
| Random (RAND) | - Benchmark<br>- Non-Adaptive | Select an unlabelled node at random. |

### A Novel Policy: Selection by Local-Entropy (LE)

One potential drawback of uncertainty-based policies is that they can lead to a bias towards selecting nodes from regions with highly heterogeneous node labels. In the context of infectious disease surveillance, this can be interpreted as an exploitation-exploration trade-off, where exploitation means the selection of nodes that lie along the boundaries between infected and uninfected regions (i.e. decision-boundaries), and exploration means the selection of nodes from less observed regions of the graph (see Fig. 2A). Previous attempts to account for this trade-off have been made, particularly in the context of AL with GNN models, whereby the exploration of less observed regions is encouraged by increasing the probability that a node is selected according to the number of unlabelled neighbours to which it is connected (44), or the degree to which the candidate node is representative of its unlabelled neighbours in feature space according to their node attributes (45). With insights from these previous efforts, we propose here a novel policy which we refer to as Selection by Local-Entropy (LE). This policy evaluates the informativeness of an unlabelled node by taking into account not only the uncertainty in the predicted label of the candidate node itself, but also that of connected nodes. At a given iteration *r*, we define the Local Entropy of an unlabelled node *v_k_* as a linear combination of the entropy of the label prediction for node *v_k_* itself, denoted by 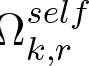, and the distance-weighted average entropy of the label predictions for surrounding nodes, denoted by 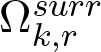, as follows,

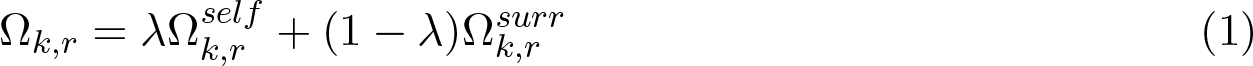

**Fig. 1.**
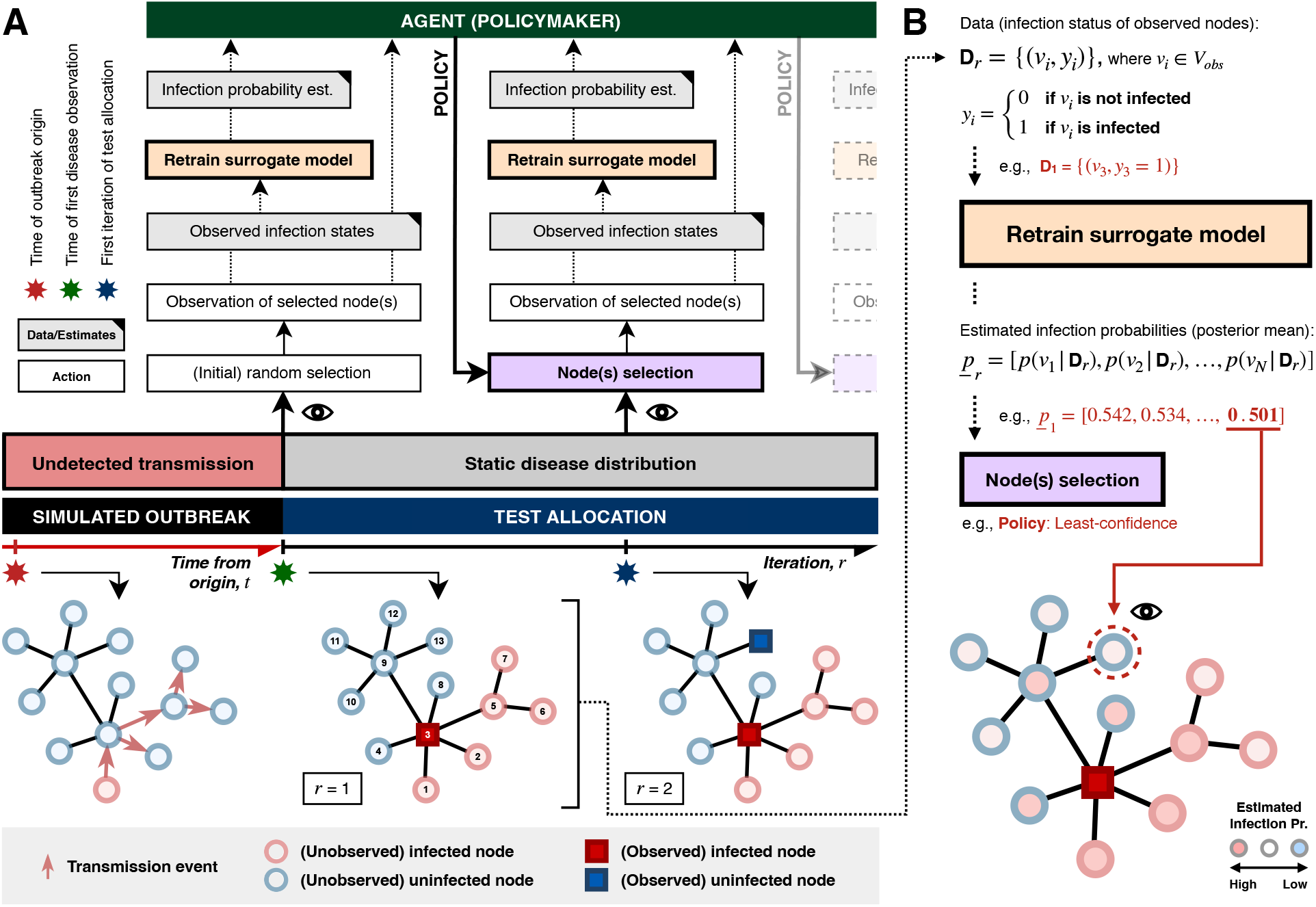
Disease surveillance on a static graph as a node classification task. (A) A schematic illustration of the adaptive test deployment framework. The flow of information/data (grey boxes with a black top-right corner) from one component (action) to another is represented as arrows. The eye symbol indicates when an observation is made, i.e. when the true infection status of a selected node is queried from the underlying disease distribution at each test iteration. Note that the simulated outbreak terminates at the time of first observation, i.e. the underlying disease distribution is assumed to be static over the course of test deployment. (B) The policy Least-Confidence (LC) in action. Information and parameter estimates specific to the example presented are highlighted in red. The initial observation of the infection status of a single node (node-3) is used as input to retrain the surrogate model, which estimates the probability of each node being infected conditional on the observed data. Given the policy LC, the node with an estimated infection probability that is closest to 0.5 is selected for observation at the next iteration (as indicated by the eye symbol). Note that node-10, -11, -12, and -13 should in principle have the same estimated infection probability given the structure of the graph, assuming a deterministic surrogate model; in such cases, one of the nodes is selected at random.

**Fig. 2.**
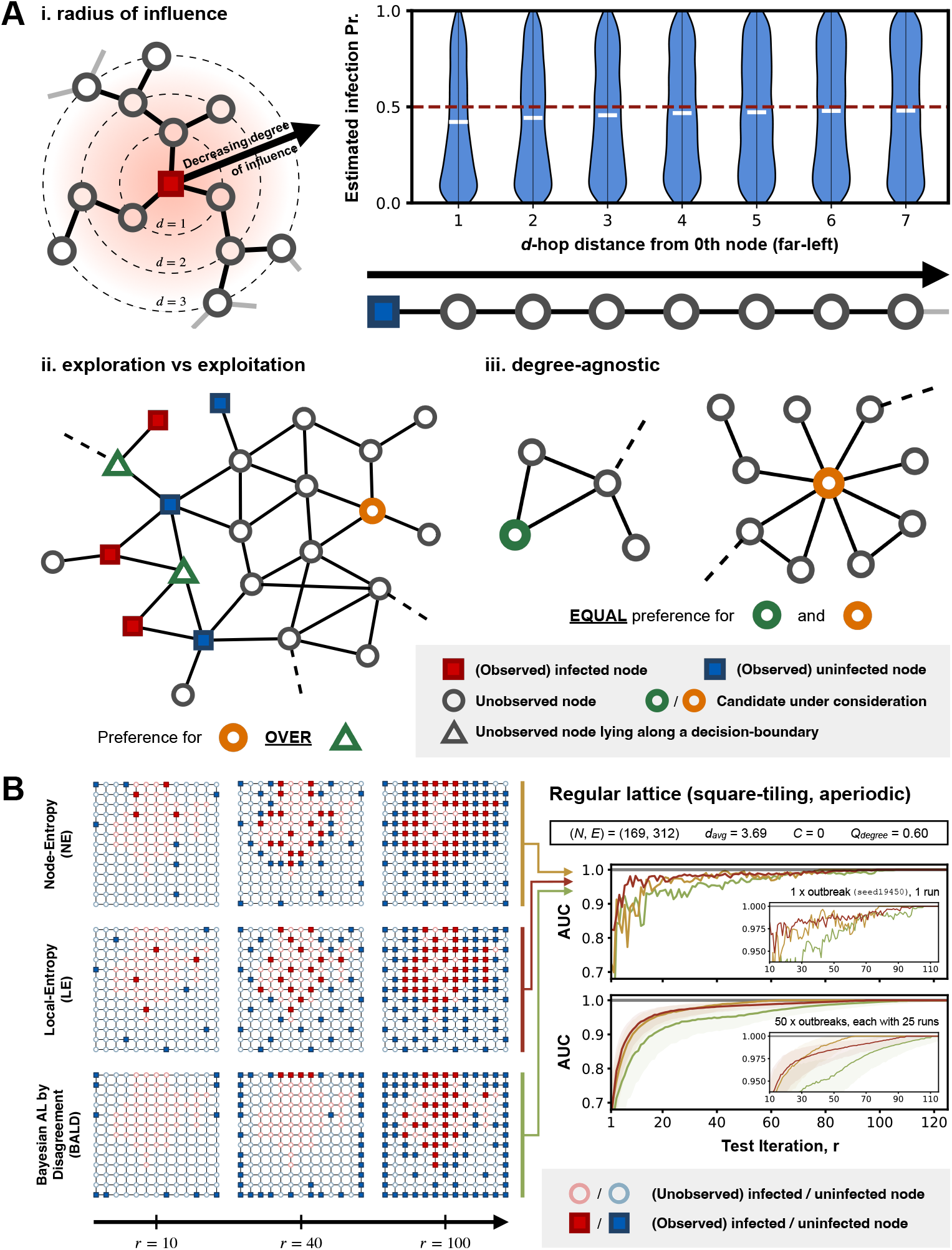
Selection by Local-Entropy (LE) and comparisons with existing uncertainty-based policies. (A) Intuitions behind the design of Selection by Local-Entropy (LE). (i, left) Schematic illustration of the decreasing degree of influence of an observation on the label prediction of surrounding nodes as the d-hop distance from the newly observed node increases. (i, right) Violin-plot of the posterior distribution of the infection probability of nodes at different d-hop distances from the only observed node (far left; node-0), arranged in a chain-like graph structure. The posterior mean of the probability of each node being infected is indicated by a white horizontal line; the red dashed line indicates a probability of 0.5 (maximally uncertain). (B) Results from a preliminary experiment in which the performance of uncertainty-based policies are compared, considering an aperiodic lattice graph with square-tiling. (left) Test allocation by three selected agents, given the same initial labelled node. Each panel shows the distribution of observed nodes (represented by coloured squares; red if infected and blue if uninfected) up to a given test iteration *r* (*r* = 10, 40, 100), as indicated by the position of the panel along the x-axis; unlabelled nodes are represented by circles, coloured according to their true infection status (red if infected and blue if uninfected, at a lower saturation compared to the observed nodes). Each row corresponds to one of the three selected agents, each assigned a different uncertainty-based policy considered in the preliminary experiment (Table 2). (right) The top plot shows the performance of the three selected agents (with each line corresponding to a different policy), as measured by the AUC. The bottom plot shows the performance of the three uncertainty-based policies, each summarised across 1250 agents (50 outbreak realisations, each with 25 unique initial labelled nodes); the shaded region represents the interquartile range and the solid line represents the median. The inset in each plot shows the same results in the interval 10 ≤ *r* ≤ 115 on an enlarged scale.

**Table 2:** Summary of all experiments conducted in this study.

| Experiment | Graph(s) | Outbreak Scenario(s) |
| --- | --- | --- |
| Preliminary<br>(*only uncertainty-based policies are considered*) | Aperiodic lattice graph (with square-tiling) | 50 random outbreak realisations, with each outbreak terminating when at least 30% of the nodes become infected ($I/N = 0.3$). |
| Synthetic Graphs | Periodic lattice graph (with square-tiling)<br>(*graph-based policies are not considered*) | 50 random outbreak realisations for each termination condition ($I/N = 0.1, 0.3, 0.5$); this amounts to a total of 150 random outbreak realisation for each graph. |
| | A random graph generated by the Barabási-Albert model, with each node having a minimum of two connections ($m = 2$) | |
|  | A random graph generated by the stochastic block model, with low-modularity settings (see S.5 in SI) |  |
|  | A random graph generated by the stochastic block model, with high-modularity settings (see S.5 in SI) |  |
| Empirical Human Mobility Networks | Graphs derived from aggregated mobility data collected from mobile phone users in Italy at provincial level during March-May, 2020 (46), with thinning-thresholds $T_{\text{thinning}} = 10\%, 15\%, 20\%$ (see S.6 in SI) | |
| | Graphs derived from global air traffic data collected at country level during Jan-March, 2020 (47), with thinning-thresholds $T_{\text{thinning}} = 2.5\%, 5\%, 7.5\%$ (see S.7 in SI) | |

with *λ* ∈ [0, 1], and

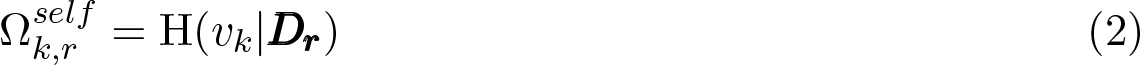

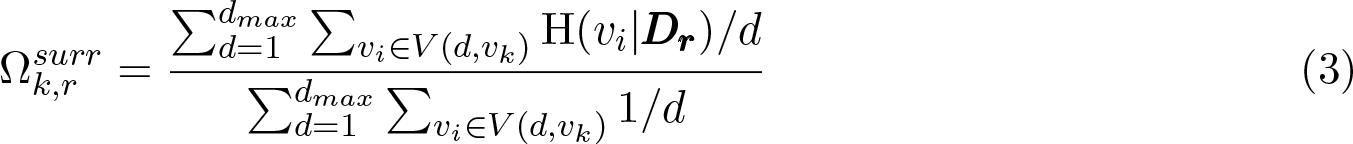

where H(*v_k_*|***D_r_***) is the entropy of the label prediction for node *v_k_*, conditional on the currently observed data ***D_r_*** = {(*v*_1_*, y*_1_), (*v*_2_*, y*_2_)*, . . .,* (*v_n_, y_n_*)}, and the entropy (for binary random variables) is defined as

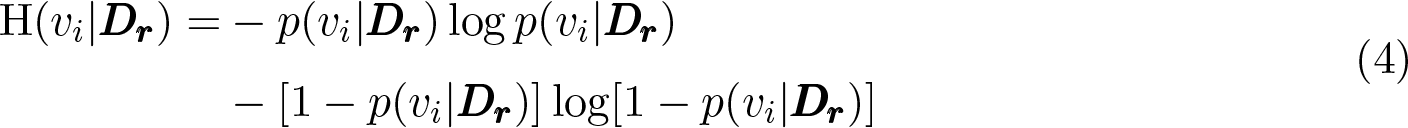

Note the double summations in the expression for 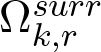 (both numerator and denominator in Equations (3)), with the first summation occurring over all neighbourhoods at different *d*-hop distances from candidate node *v_k_* for *d* ∈ [1*, d*_max_]. Here, *d*_max_ is an integer parameter whose value is bounded by the diameter of the graph, *d_G_*, i.e. the greatest geodesic distance between any pair of nodes; it determines the cut-off in *d*-hop distance beyond which the observed label of a node is assumed to have a negligible effect on the label prediction of an unobserved node (radius of influence; Fig. 2A, top-left). The second summation occurs over the entropy of the label prediction for all nodes in a given *d*-hop neighbourhood of the candidate node *v_k_* (denoted by *V* (*d, v_k_*)), weighted by the inverse of the geodesic distance *d*.

Key insights that motivate the definition of Local Entropy can be summarised as follows:

1. The information that can be gained from the observation of a node is likely to be greater if it is in close proximity to other unlabelled nodes with highly uncertain label predictions.
2. The influence that a new observation has on the label prediction of surrounding nodes decays with increasing *d*-hop distance. This, together with insight (1), motivates the definition of 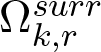, i.e. the sum of the entropy of the label prediction of all surrounding nodes (up to *d*-hop distance *d_max_*) weighted by 1*/d*, as a proxy measure of the total impact that the new observation is likely to have on the label predictions of surrounding nodes.
3. This sum, as described in (2), is normalised by sum of the distance-weights across all *d*-hop neighbourhoods (up to *d*-hop distance *d_max_*); this is to avoid the bias where centrally located nodes would have larger values of 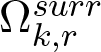, simply as a result of having more connections.
4. The balance between exploitation and exploration (see above) can be fine-tuned by specifying different values of *λ*. In the case where *λ* = 1, we recover the uncertainty-based policy which performs node selection based on node-entropy alone.

Note that we set *d*_max_ to the graph diameter, *d_G_*, in all our following experiments; we leave the exploration of different values of *d*_max_ for future work. We also set *λ* = 0 in all subsequent considerations of our proposed policy LE (i.e. maximal exploration).

### Synthetic Graphs and Empirical Human Mobility Networks

To understand better how the policies perform under different outbreak scenarios and on networks with different structural properties, we conduct three sets of experiments (Table 2). The different graph models were chosen to ensure a diverse range of degree distributions and varying levels of community structure and structural disorder.

For each random outbreak realisation on a given network, 25 different nodes are randomly selected as the initial labelled node; at the beginning of each experiment, the infection status of the same initial labelled node is made available to all agents to ensure a fair comparison between policies. This is done to account for any variability in policy performance resulting from different initial observations, stochasticity from the Markov chain Monte Carlo (MCMC) inference process, or from random tie-breaking whenever two or more candidate nodes are given equal preference by a policy according to its selection criterion.

In the experiment where we consider empirical human mobility data (Table 2, row 3), an unweighted and undirected graph is constructed from each dataset by (i) first symmetrising the matrices representing the mobility flows and (ii) averaging them over the collection period, followed by (iii) removing edges with mobility flow below a certain threshold (also known as graph-thinning; see S.6 and S.7 in SI for a detailed description). We perform a series of sensitivity analyses to ensure that our results are robust to different thinning-thresholds (see Fig. S3 and S4 in SI).

### Test Budget and Performance Evaluation

The performance of each policy at a given iteration *r* is evaluated by calculating the Area Under the Receiver Operating Characteristics Curve (AUC), based on the model predictions from the surrogate model, conditional on the observed data up to the current iteration *r*. An AUC of 0.5 indicates no discriminative power and 1 indicates perfect predictions. Note that the AUC is only evaluated for the set of nodes with unknown infection status at each iteration. This is because, in the context of disease surveillance, the performance of an agent is ultimately determined by the degree to which it is able to predict the infection status of unobserved locations, with the distribution of these unobserved locations in the network in turn determined by the allocation policy of choice.

We also consider a policy referred to as Reactive-Infected (RI) designed to mimic the decisions of a policymaker whose aim is to identify as many infected locations as possible with the given resources, i.e. a “contact-tracing” approach. This policy provides a benchmark for the average test budget required to identify all infected nodes in a given outbreak scenario. It is therefore only at test iterations below this test budget that the objective of accurately predicting the presence or absence of a disease of interest may be considered relevant to public health decisions. In all following experiments, we compare the performance of the different policies only at test iterations up to this benchmark (median number of test iterations needed by RI to identify all infected nodes across all simulated outbreak scenarios for a given graph); full results are presented in Fig. S1 and S2 in SI.

## Main Results

### Disease Surveillance on an Aperiodic Regular Lattice Graph

As a preliminary experiment to illustrate the differences between the uncertainty-based policies considered, we evaluate and compare their performance on an aperiodic regular lattice graph with square-tiling. We observe that our proposed policy LE on average performs better than both NE and BALD at small numbers of test iterations (*r <* 30; Fig. 2B, bottom-left). LE and NE show similar performance between *r* = 30 and *r* = 50; at *r >* 50, however, NE overtakes LE as the best performing policy with an AUC that rapidly approaches 1, while both LE and BALD struggle to attain a perfect AUC. The difference in performance between LE and NE can be understood in the context of the exploitation-exploration trade-off, as described previously: at small *r*, LE encourages an even allocation of tests across the graph (exploration), while NE favours regions with highly heterogeneous disease distributions (exploitation) (see Fig. 2B, first and second row on the left) - this results in a more rapid increase in model performance for LE as *r* increases. At large *r*, however, the greater preference for exploitation by NE results in almost all of the nodes that lie along the decision-boundary being sampled - this results in an AUC that rapidly approaches 1. Although LE also shows a preferential selection of nodes close to the decision-boundary at large *r*, it does so at a much slower rate compared to NE.

BALD (40) performs on average worse than both NE and LE across all test iterations. This is due to its apparent preferential selection of low-degree nodes (either in the corners or along the edges); only at *r >* 40 (at which point no low-degree nodes remain) do we see a pattern of test allocation that resembles that of NE. The observed underperformance of BALD is consistent with results from a previous comprehensive evaluation of existing AL policies for node classification (48), likely explained by the fact that BALD does not take into account the graph structure in its formulation (40).

### Disease Surveillance on Synthetic Graphs

There are three key observations from our results presented in Fig. 3. First, all policies except for BALD and RI outperform random allocation (RAND) across all outbreak scenarios, especially at large *r* when the performance of random allocation appears to only increase slowly with increasing *r*. Given the preferential selection of low-degree nodes by BALD as described in the previous section, it is not surprising that BALD only shows comparable performance in the periodic lattice graph which has no degree variation. Secondly, uncertainty-based policies (NE, BALD and LE) underperform substantially compared to graph-based heuristics (DC, PC) on the synthetic graph generated by the Barabási-Albert model (hereafter referred to as the BA-graph). This observation can be explained by considering the infection-assortativity, *r*_infection_, which in the context of disease distribution, is a measure of the tendency for two connected nodes to share the same infection status (as has been repeatedly shown in empirical studies that mobility synchronises epidemics across locations (49); see S.8 in SI for definition of infection-assortativity). Evaluating the average *r*_infection_ value across all outbreak realisations on each graph shows that outbreaks on the BA-graph have on average the lowest *r*_infection_ at 0.20 (compared to 0.64 for the periodic lattice graph, 0.48 and 0.63 for the graphs generated by the stochastic block model (SB-graph) with low and high modularity (50), respectively). A low (but positive) *r*_infection_ value indicates a weak tendency for two connected locations to share the same infection status, and therefore a low degree of homophily in the underlying disease distribution. This results in an overall poor predictive performance from the surrogate model, which in turn limits the effectiveness of the uncertainty-based policies. In such cases, it may then be advantageous to consider node-centrality alone during node selection, especially at small *r* when there is little data to inform model predictions. Note also that PC tends to perform better than DC - this is not unexpected given that nodes with the most connections are not necessarily the most central in a network.

**Fig. 3.**
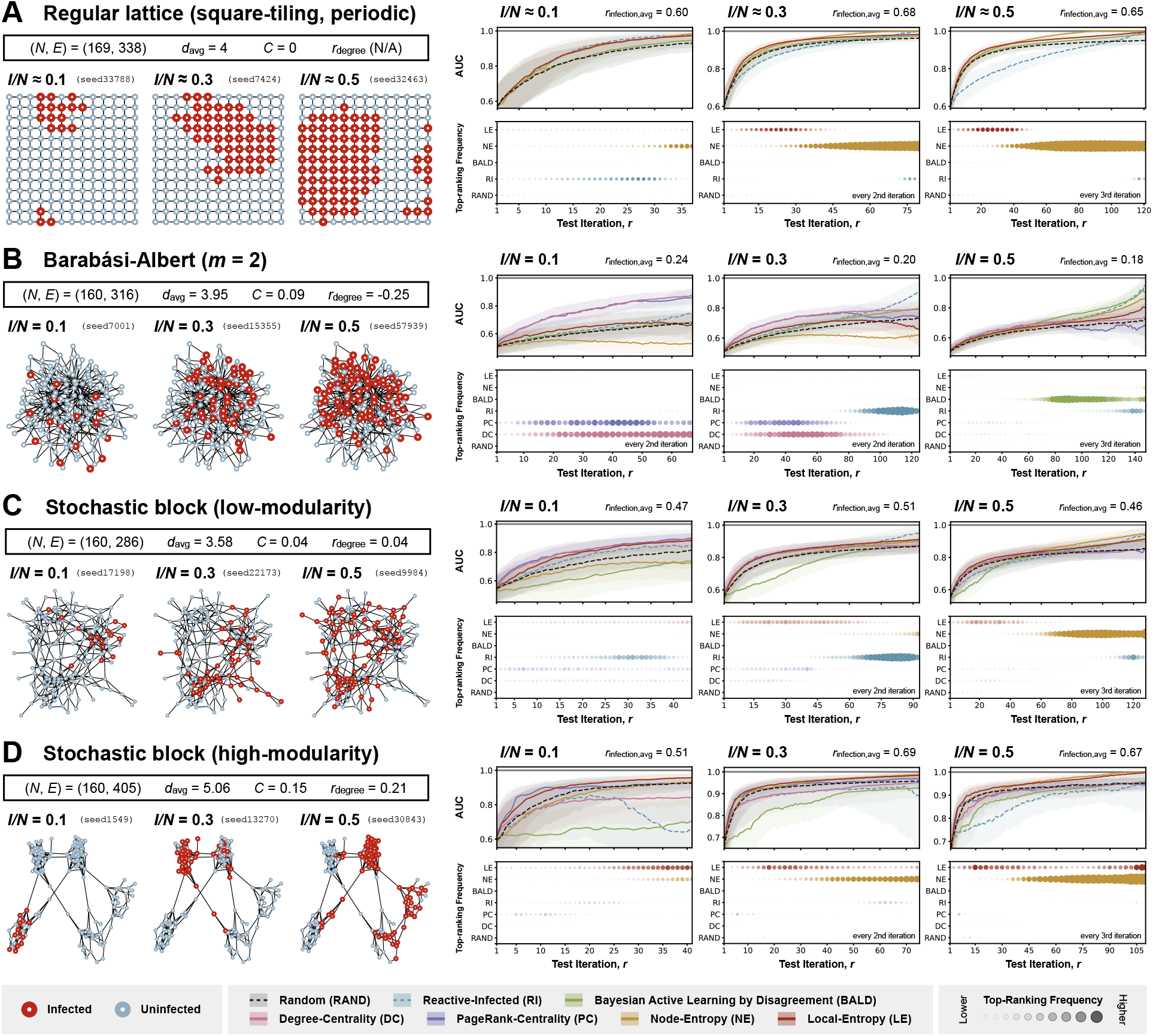
Policy evaluation with simulated outbreaks on synthetic graphs. Each panel (A, B, C, and D) corresponds to results from experiments with simulated outbreaks on a different synthetic graph (panel A: a periodic regular lattice graph with square-tiling; panel B: a random graph generated by the Barabási-Albert model, with each node having a minimum of two connections (*m* = 2); C: a random graph generated by the stochastic block model at low-modularity settings; D: a random graph generated by the stochastic block model at high-modularity settings). Summary statistics relevant to the structure of each graph (number of nodes (*N*), number of edges (*E*), average node-degree (*d*_avg_), clustering coefficient (*C*), and degree-assortativity (*r*_degree_)) are shown in the top-left part of each panel. In the bottom-left part of each panel are visualisations of three selected disease distributions (with their corresponding seeds shown), each at a different stage of outbreak progression as measured by the proportion of nodes infected (*I/N* = 0.1, 0.3, 0.5); nodes are coloured according to their true infection status (red if infected and blue if uninfected). In the right part of each panel, each column shows results from experiments considering disease distributions at a different stage of outbreak progression. The top plot in each column shows the performance of policies considered in the corresponding experiment, as measured by the AUC, with a higher AUC indicating a better performance; the shaded region represents the interquartile range and the solid line represents the median. The bottom plot in each column shows the frequency with which each policy is ranked top according to its AUC at each iteration (or every 2nd or 3rd iteration, as indicated); a larger circle with a more saturated colour indicates a higher frequency of the policy being ranked top at a given iteration. The performance of each policy is only shown up to the median number of test iterations required for all infected nodes to be observed among agents assigned to Reactive-Infected (RI) (see Fig. S1 in SI for full results).

Finally, we observe generally favourable performance for LE across most of the outbreak scenarios on graphs with a high degree of structural order (unlike the BA-graph, as described), especially at small *r*. At larger *r*, however, we again observe superior performance for NE, with AUCs that rapidly approach 1. This can again be explained by the preference for exploitation over exploration by NE, which leads to the complete observation of the decision-boundary between infected and uninfected regions given a sufficient number of test iterations.

### Disease Surveillance on Empirical Human Mobility Networks

From Fig. 4, it is evident that the two graphs derived from empirical human mobility data have structural properties that are markedly different from each other. Graph A, generated from an openly available aggregated mobility data derived from mobile phone trajectories in Italy at provincial level (46), shows distinct community structures with close resemblance to the SB-graphs as described in the previous section. In contrast, Graph B, generated from the global air traffic data collected at country level (47), displays structural properties that are similar to those of the BA-graph. This is consistent with previous studies that show the global air traffic network has scale-free properties (51, 52) (e.g., both have a negative degree-assortativity (see Fig. 4B), indicating a hub-and-spoke as opposed to hub-and-hub structure (53)).

**Fig. 4.**
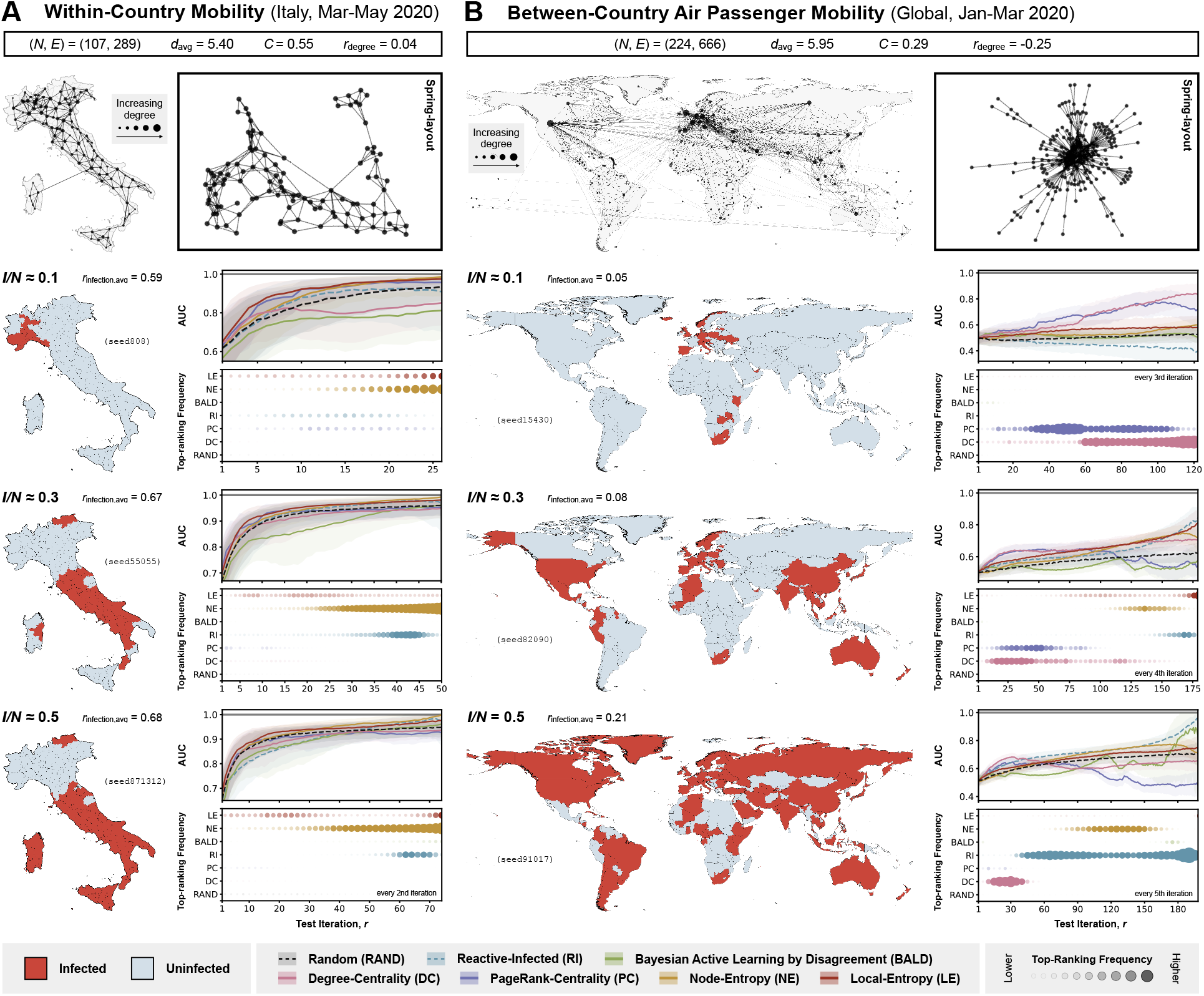
Policy evaluation with simulated outbreaks on graphs derived from empirical human mobility data. Each panel (A and B) corresponds to results from experiments with simulated outbreaks on a graph derived from a different empirical human mobility dataset (panel A: within-country movements from smartphone data collected in Italy at provincial level between March-May 2020 (46), with thinning-threshold *T*_thinning_ = 15%; panel B: between-country human movement from air traffic data collected between January-March 2020 (47), with thinning-threshold *T*_thinning_ = 5%). Summary statistics relevant to the structure of each graph (number of nodes (*N*), number of edges (*E*), average node-degree (*d*avg), clustering coefficient (*C*), and degree-assortativity (*r*_degree_)) are shown at the top of each panel, followed by (left) a visualisation of the graph overlaid on a corresponding map (with the size of each node indicating the node-degree) and (right) a visualisation of the same graph in spring-layout. The 2nd to 4th row of each panel correspond to the different stages of outbreak progression at which the performance of the different policies are evaluated, as measured by the proportion of nodes infected (*I/N* = 0.1, 0.3, 0.5). In the left part of each row is the visualisation of a selected disease distribution (from one of 50) on the corresponding map; tiles are coloured according to their true infection status (red if infected and blue if uninfected). In the right part of each panel, each column shows results from experiments considering disease distribution at a different stage of outbreak progression. The top plot in each column shows the performance of policies considered in the corresponding experiment, as measured by the AUC, with a higher AUC indicating a better performance; the shaded region represents the interquartile range and the solid line represents the median. The bottom plot in each column shows the frequency with which each policy is ranked top according to its AUC at each iteration (or every 2nd or 3rd iteration, as indicated); a larger circle with a more saturated colour indicates a higher frequency of the policy being ranked top at a given iteration. The performance of each policy is only shown up to the median number of test iterations required for all infected nodes to be observed among agents assigned to Reactive-Infected (RI) (see Fig. S2 in SI for full results).

We observe policy performances on Graph A and Graph B that are similar to those from experiments on the SB-graphs and BA-graph, respectively. Most notably for Graph A, LE again shows rapid increases in model performance given small numbers of test iterations, only to be surpassed by NE at large *r*, as expected. For Graph B, graph-based policies (DC, PC) outperform uncertainty-based policies especially at small *r*, again consistent with results from experiments on the BA-graph. However, the superior performance of these graph-based policies only extends to larger values of *r* if the outbreak under surveillance is in its early stages (i.e. *I/N* = 0.1); at later stages of outbreak progression, the performance of these policies decreases with further increases in *r*.

This counterintuitive observation can be explained by considering the changes in the distribution of the decision-boundary between the infected and uninfected regions in the graph during a transmission process. At the beginning of an outbreak, nodes that are centrally located are more likely to be infected early on due to their high degree of connectivity. This implies that most of the decision-boundary between infected and uninfected regions can be found close to the central nodes, thus explaining the superior performance of graph-based policies which preferentially select nodes with a high degree of centrality. As the outbreak progresses, the decision-boundary shifts towards the periphery of the graph with the already infected central nodes acting as secondary hubs of the emerging pathogen. This results in a decrease in the performance of graph-based policies, as the central nodes continue to be targeted while the peripheral regions of the graph (where most heterogeneities in the disease distribution lie) remain largely unexplored. Note that a similar drop in the performance of PC (Fig. 3B, second and third column) at large *r* during later stages of outbreak progression (*I/N* = 0.3 and *I/N* = 0.5) can also be observed.

The same reasoning can also potentially explain the unexpected superior performance of graph-based policies and comparable performance of RI at small *r*, especially during the early stage of an outbreak (see Fig. 3). More generally, provided that the number of infected nodes is sufficiently small and that they are confined to a small, local region of the graph, any policy for which there is a high probability of selecting an infected node is likely to perform well compared to other policies, especially when given a small test budget.

## Discussion

In this work, we addressed the question of how a finite amount of testing resources should be allocated across a network of locations connected by mobility in order to maximise the information gained about the underlying distribution of an infectious disease. We formulate this task as a node classification problem with AL, with the objective of providing accurate assessment of where the disease is likely to be present or absent given a fixed test budget. We proposed a novel policy which selects nodes for observation according to a measure of the distanced-weighted average entropy of the label predictions in the local neighbourhood of a given candidate node. We then evaluated and compared the performance of different policies, including our proposed policy, under a range of different outbreak scenarios and graph structures.

Our results show that in general there is not a single policy that performs optimally across all outbreak scenarios. Instead, the performance of a given policy depends on both the test budget available relative to the size of the network and the geometry of the underlying disease distribution, which is in turn determined by the network structure and the stage of the outbreak. For example, graph-based policies that target central nodes perform better than uncertainty-based policies when the underlying disease spread cannot be modelled with a high degree of accuracy and certainty, as is often the case during early stages of an outbreak when the aetiology is still unknown. Conversely, uncertainty-based policies are typically more effective in highly ordered networks with well-defined community structures. In particular, with our proposed policy (Local-Entropy) which considers graph-based uncertainties in its selection criterion, we were able to show that more frequent exploration results in better performance given a small test budget, while targeting regions in the network with observed heterogeneous disease distribution (exploitation) is more favourable given a large test budget. Finally, we find that following an approach akin to contact-tracing (selecting neighbours of infected nodes) generally leads to inferior performance compared to other policies in terms of characterising the overall disease distribution. A comprehensive assessment of the overall distribution could potentially allow for a more detailed study of the underlying transmission process (e.g., identifying drivers of spread by iteratively re-fitting prediction models of disease progression on a network), and provide an opportunity to improve the joint modelling of infectious diseases and sampling more generally.

Although we observe consistent results across experiments on both synthetic graphs and empirically-derived networks, it is important to interpret these findings in the context of the assumptions made, particularly regarding their generalisability to real-world scenarios. One important limitation of our approach is the assumption that the underlying mobility network is represented as an undirected and unweighted graph. In practice, real-world mobility networks are highly heterogeneous and have mobility fluxes that vary across both regions (e.g., air traffic among European countries versus African countries) and directions (e.g., net inflow of air passengers arriving at tourist destinations during holiday seasons). Future extensions should consider alternative surrogate models that are able to incorporate these effects when generating label predictions, e.g., Gaussian Processes on graphs and spatial mechanistic models that explicitly model the movement of infectious individuals. Another limitation of this study is the assumption of static disease distributions. This implies that the timescale over which transmission events between locations occur is sufficiently longer than the timescale of test deployment, such that the underlying disease distribution can be treated as static. While this is unlikely to be a realistic assumption for most disease outbreaks - except for some endemic diseases that are more slowly-changing in their prevalence, such as HIV/AIDS (54) and Tuberculosis (TB) (55) - it nevertheless allowed us to gain theoretical insights on the various considerations one must take when designing disease surveillance strategies given different network structures and outbreak scenarios. To address this limitation, future work should consider the correlation in infection status not only between nodes but also across time, given either prior assumptions of the underlying transmission dynamics or information from historical transmission events that are inferred to have occurred given the data. In this dynamic setting, it might also be advisable to consider testing multiple locations at once (similar to batch AL (56), as opposed to only a single location per iteration as presented in our work. Finally, we assume that there is no observational noise, i.e. the true infection status of a selected node is revealed upon testing at each iteration - future studies should consider the impact of misclassification and therefore the need to deploy multiple tests depending on both test sensitivity and the estimated prevalence at a given location.

Our findings in this study are particularly relevant to infectious disease surveillance in resource-constrained settings and in situations where practical challenges render the complete detection of all infected populations unfeasible or cost-inefficient. Here we propose a flexible and principled approach to evaluating the design and execution of adaptive surveillance strategies with the overall aim of maximising the information gained from each test iteration. More generally, our adaptive test deployment framework can be extended to consider transmission processes with greater complexities (e.g., SEIR models, spatially-explicit semi-mechanistic models) and more realistic mobility networks (e.g., as directed and weighted graphs, with time-varying edge weights) that are derived from empirical data, with additional constraints to account for imperfect testing (e.g., observational noise and delay in test feedback). Applications of our model in real-world contexts could provide the opportunity for more cost-effective and rapid identification and monitoring of pathogens while reducing the uncertainties associated with early risk assessments of infectious diseases.

## Data, Materials, and Software Availability

All data, code and analysis files used in this work will be made openly accessible from GitHub at https://github.com/joetsui1994/optimal-disease-surveillance-AL.

## Data Availability

All data produced in the present study will be made available online at https://github.com/joetsui1994/optimal-disease-surveillance-AL

## Acknowledgements

M.U.G.K. acknowledges funding from The Rockefeller Foundation, Google.org, the Oxford Martin School Pandemic Genomics programme (also O.G.P. & J.L.-H.T.), European Union’s Horizon Europe programme projects MOOD (#874850) and E4Warning (#101086640), the John Fell Fund, a Branco Weiss Fellowship and Wellcome Trust grants 225288/Z/22/Z, 226052/Z/22/Z & 228186/Z/23/Z, United Kingdom Research and Innovation (#APP8583) and the Medical Research Foundation (MRF-RG-ICCH-2022-100069). J.L.-H.T. is supported by a Yeotown Scholarship from New College, University of Oxford. The contents of this publication are the sole responsibility of the authors and do not necessarily reflect the views of the European Commission or the other funders. E.S. acknowledges support in part by the AI2050 program at Schmidt Futures (Grant [G-22-64476]). M.A.S. is supported in part through the US National Institutes of Health grant R01 AI153044. S.F. and M.Z. acknowledge the EPSRC (EP/V002910/2).

