## Supplementary Information for "Optimal disease surveillance with graph-based Active Learning"

#### S.1 Stochastic Susceptible-Infected Process on Graphs

Each node is initially in the susceptible (S) state. At $t = 0$, a single node is randomly selected and set to the infected (I) state. At each following time step where $t > 0$, we assume that there is a fixed probability $p = 0.1$ for any infected node to infect their susceptible neighbours, i.e. any nodes that are in the susceptible state and are connected to the infected node by an edge. In this simple SI process, it is assumed that infected nodes do not recover or become immune - once infected, they remain infected indefinitely with the same constant probability of onward transmission, $p$, throughout the rest of the simulation. This process continues until a certain proportion of nodes are infected, as specified by a target $I/N$ value, where $I$ is the number infected nodes and $N$ is the total number of nodes in the graph. Different values of $I/N$ indicates different stages of outbreak progression at the time of disease surveillance.

An important implication resulting from the assumptions made in the SI process as described, is that a node can only be infected if at least one of its immediate neighbours is also infected (with the exception of the initially infected node), i.e. all infected nodes must be connected in the graph. This implies that there can only be a single infected region, however with potentially multiple uninfected regions and therefore multiple decision-boundaries (lines or surfaces separating infected and uninfected regions). The distribution of these decision-boundaries vary between outbreaks depending on both the network structure and the stage of outbreak progression (proportion of nodes that are infected).

#### S.2 Conditional Autoregressive Model (CAR) as A Surrogate Model

The Conditional Autoregressive (CAR) model (1) is widely used in the small area estimation domain, where data consist of a set of observations $\boldsymbol{y} = [y_1, y_2, \ldots, y_n]$ over a set of $n$ spatial units, which in the context of our study represent locations in a mobility network. The CAR model assumes that the value of a variable in one location (node, or location) depends on the values of neighbouring locations, with weights specified by a spatial adjacency matrix $\boldsymbol{A}$. For unweighted models, like the one we are working with in this paper, the adjacency matrix $\boldsymbol{A}$ is binary and captures the presence or absence of edges between corresponding nodes. The spatial random effect $\boldsymbol{f} = [f_1, f_2, \ldots, f_n]$ follows the multivariate normal prior with precision matrix $\boldsymbol{Q}$:

$$\boldsymbol{f} \sim \mathcal{N}(0, \boldsymbol{Q}^{-1}) \tag{1}$$

$$\boldsymbol{Q} = \tau(\boldsymbol{A} - \alpha\boldsymbol{D}) \tag{2}$$

The parameter $\alpha$ captures the amount of spatial correlation between connected nodes, and can take any value between 0 and 1 (inclusive). If $\alpha = 0$, the model reduces to a set of independent errors at every location, and if $\alpha = 1$, the model reduces to the ICAR (intrinsic conditional autoregressive model) - another, but less flexible model. In this study, we set a fixed value for $\alpha$ with $\alpha = 0.95$ to clearly separate the tasks of spatial inference on graph from the task of optimisation and use $\tau \sim \text{logNormal}(0, 0.1)$ as prior on the marginal precision.

CAR, as well as ICAR, are standard models in spatial statistics. Similarly as Gaussian Processes (GPs) are a standard choice for surrogates over continuous space, CAR is the default model choice for modelling over a discrete set of areas. Future work includes a wider range of surrogates, such as GPs on graphs when no knowledge about the spread of the disease is available, or mechanistic models, such as SIR, SEIR and similar, when it is reasonable to make an assumption about the mechanics of the disease spread.

#### S.3 Bayesian Active Learning by Disagreement (BALD)

Bayesian Active Learning by Disagreement (BALD) (2) is one of the state-of-the-art acquisition policies in Active Learning. It selects the data instances that maximise the decrease in expected posterior entropy,

$$v_{r+1} = \underset{v \in V}{\text{argmax}}\, \mathrm{I}(\theta; y|v, \boldsymbol{D_r}) \tag{3}$$

where $\theta$ is the latest parameters and $\boldsymbol{D_r}$ is the set of data instances labelled up iteration $r$, and the mutual information, $I$, is defined as follows:

$$\begin{aligned} \mathrm{I}(\theta; y|v, \boldsymbol{D_r}) &= \mathrm{H}(\theta|\boldsymbol{D_r}) - E_{y \sim p(y|v, \boldsymbol{D_r})} \mathrm{H}(\theta|y, v, \boldsymbol{D_r}) \\ &= \mathrm{H}(y|v, \boldsymbol{D_r}) - E_{\theta \sim p(\theta|\boldsymbol{D_r})} \mathrm{H}(y|\theta, v, \boldsymbol{D_r}) \\ &= \mathrm{H}\left[\int p(y|v, \theta) p(\theta|\boldsymbol{D_r}) d\theta\right] - \int \mathrm{H}[p(y|v, \theta)] p(\theta|\boldsymbol{D_r}) d\theta \\ &\approx \mathrm{H}\left[\frac{1}{n}\sum_{i=1}^{n} p(y|v, \theta_i)\right] - \frac{1}{n}\sum_{i=1}^{n} \mathrm{H}[p(y|v, \theta_i)] \end{aligned} \tag{4}$$

where $\theta_i \sim p(\theta|\boldsymbol{D_r})$.

For Gaussian Process classification tasks, Houlsby et al. (2011) (2) provided approximations of BALD. This formulation highlights that the mutual information can be approximated using posteriors obtained numerically. Hence, one can use surrogates of any complexity as long as their parameters can be estimated in a Bayesian manner.

#### S.4 Selection by Least-Confidence (LC) and Node-Entropy (NE)

The policy Least-Confidence (LC) selects the node for which the surrogate model is the least confident about its label prediction (label with the highest estimated probability among possible labels). More formally, this can be written as

$$v_{r+1} = \arg\min_{v \in V} \max_{y \in Y} p(y|v, \boldsymbol{D_r}) \tag{5}$$

where $Y$ is the set of possible labels, $V$ is the set of unlabelled nodes available for selection, and $p(y|v, \boldsymbol{D_r})$ is the estimated probability of node $v$ having label $y$, conditioned on the observed data up to iteration $r$, $\boldsymbol{D_r}$. In the special case of binary classification where $Y \in \{0, 1\}$, since the most likely label must by definition have an estimate probability greater than 0.5, this policy reduces to selecting the node with an estimated probability of having either label that is closest to 0.5. Without loss of generality, let $p(v|\boldsymbol{D_r})$ denote the probability of node $v$ having label $y = 1$ conditioned on the observed data up to iteration $r$, the policy LC can then be written as

$$v_{r+1} = \arg\min_{v \in V} |p(y|v, \boldsymbol{D_r}) - 0.5| \tag{6}$$

The policy NE selects the node with the highest entropy in predicted label distribution, i.e.

$$v_{r+1} = \arg\min_{v \in V} \mathrm{H}[p(y|v, \boldsymbol{D_r})] \tag{7}$$

where

$$\mathrm{H}[p(y|v, \boldsymbol{D_r})] = -\sum_{y \in Y} p(y|v, \boldsymbol{D_r}) \log p(y|v, \boldsymbol{D_r}) \tag{8}$$

In the special case of binary classification, this expression reduces down to

$$\begin{aligned} \mathrm{H}[p(y|v, \boldsymbol{D_r})] = \mathrm{H}[p(v|\boldsymbol{D_r})] &= -p(y|v, \boldsymbol{D_r}) \log p(y|v, \boldsymbol{D_r}) \\ &\quad - [1 - p(y|v, \boldsymbol{D_r})] \log[1 - p(y|v, \boldsymbol{D_r})] \end{aligned} \tag{9}$$

with $p(v|\boldsymbol{D_r})$, again, being the probability of node $v$ having label $y = 1$, conditioned on the observed data up to iteration $r$.

From (9), it is straightforward to see that $\mathrm{H}[p(v|\boldsymbol{D_r})]$ is a concave function of $p(v|\boldsymbol{D_r})$ that is symmetric about the line $p(v|\boldsymbol{D_r}) = 0.5$, i.e. when there is equal probability for node $v$ to have either label. As a result, the node with an estimated probability $p(v|\boldsymbol{D_r})$ that is closest to 0.5 must also be the node with the highest entropy $\mathrm{H}[p(v|\boldsymbol{D_r})]$. Therefore, in the special case of binary classification, the policy LC always selects the same node as the policy at NE at each iteration.

### S.5 Generating Random Graphs with Community Structure Using the Stochastic Block Model

We used the stochastic block (SB) model to generate random graphs with different levels of community structure. We began by first specifying the number of communities, $k$, and the size of each community. In this study, we set $k = 5$ with the size of each community selected at random while keeping the total number of nodes in the graph at 160. To control the level of community structure, we varied the value of the parameter $p_{\text{intra}}$ and $p_{\text{inter}}$, i.e. the probability of connection within a community and between communities, respectively. For example, a high $p_{\text{intra}}$ with a low $p_{\text{inter}}$ indicates a strong community structure, with nodes within communities being tightly connected and only sparse connections between communities. To generate a random graph with a high level of community structure, we set the parameter $(p_{\text{intra}}, p_{\text{inter}}) = (0.14, 0.001)$; and to generate a random graph with a lower of community structure, wet set $(p_{\text{intra}}, p_{\text{inter}}) = (0.08, 0.005)$.

One common way to quantify the level of community structure present in a graph is to compute its modularity [31]. The modularity of a graph is a measure of the degree to which it can be partitioned into distinct modules or communities; it is defined as the fraction of edges that fall within communities minus the expected fraction of edges that would fall within communities if edges are distributed randomly. Given a graph with adjacency matrix $\boldsymbol{A}$, its modularity is given by

$$Q = \frac{1}{2m} \sum_{ij} \left[ \boldsymbol{A}_{ij} - \frac{k_i k_j}{2m} \right] \delta(c_i, c_j) \tag{10}$$

where $m$ is the total number of edges in the graph, $k_i$ and $k_j$ are the degrees of node $i$ and $j$, and $\delta(c_i, c_j)$ is 1 if node $i$ and $j$ are in the same community, and 0 otherwise.

Applying the above formula shows that the random graph generated using the SB model with a high level of community structure has a modularity of 0.72, whereas the random graph with a lower level of community structure has a modularity of 0.62.

#### S.6 Pre-Processing of Within-Country Human Mobility Data Collected at Provincial Level in Italy

A dataset containing daily aggregated mobility data collected from mobile phone users in Italy at provincial level, covering the period between 18 January and 26 June 2020 (3), was downloaded from `https://covid19mm.github.io/data.html` on 26 February 2024. The data consists of the daily number of smartphone users moving both within and between 107 provinces, normalised by the number of active users each week, which has been shown to be roughly constant throughout the collection period (3). Here we focus our analysis on the period between March and May (inclusive), during which a national lockdown (from 9 March to 18 May, 2929) was imposed by the Italian government in response to the emerging COVID-19 outbreak.

To construct a static graph from the mobility data, we first summed the mobility flows over both directions for each pair of provinces to obtain a symmetric matrix for each day, which was then averaged across the 6-month period. The resulting matrix was then converted into an unweighted graph using a procedure known as graph-thinning. In this process, edges representing pairs of provinces were ranked according to their total mobility flow as calculated earlier; edges were then removed one at a time starting from those ranked the lowest while ensuring that the graph remained connected. This iterative process continued until a certain target proportion of the original edges remained; this target proportion is known as the thinning-threshold. Finally, the all edge weights are removed.

The choice of this threshold takes into consideration the balance between 1) the need to remove edges with very low mobility flows and are therefore less relevant to the overall structure of the graph, versus 2) the need to retain enough edges in order to preserve important structural properties (e.g., presence of travel hubs and community structure) of the graph. With these in mind, the thinning-thresholds of 10%, 15% and 20% were specified. To ensure robustness, the same experiments were repeated on each of the resulting graphs (see Fig. S3); however, only results from experiments performed on the graph with a thinning-threshold of 15% are presented in Fig. 4.

#### S.7 Pre-Processing of Between-Country Air Traffic Data Collected at Country Level

A dataset containing monthly air traffic data collected at country level, covering the period between January and March 2020 (4), was downloaded from `https://zenodo.org/records/7472836` on 1 March 2024. The data consists of the monthly number of air passengers travelling both within and between countries. To construct an undirected and unweighted graph from the data, the same procedure as described in S.6 was performed. Due to the much greater number of edges (as a result of a greater number of nodes and the presence of long-range movements in the air traffic network), a lower thinning-threshold was used to ensure the surrogate model can be fitted within a reasonable timeframe at each iteration given the available computational resources. With the considerations as described in S.6, the thinning-thresholds of 2.5%, 5% and 7.5% were specified. Again, the same experiments were repeated on each of the resulting graphs to ensure robustness of our results (see Fig. S4); however, only results from experiments performed on the graph with a thinning-threshold of 5% are presented in Fig. 4.

#### S.8 Degree-Assortativity and Infection-Assortativity

Degree-assortativity of a network, commonly denoted as $r_{\text{degree}}$, is a measure of the tendency for nodes to connect with other nodes with similar degrees. It can take any value between -1 and 1, with a positive value indicating that high-degree nodes are more likely to connect with other high-degree nodes, and similarly for low-degree nodes (assortative mixing by degree). Conversely, a negative value indicates a tendency for high-degree nodes to connect with low-degree nodes, and vice versa (disassortative mixing by degree).

The same idea of assortativity can be extended to other node attributes, including infection status as considered in this study. A positive assortativity by infection status (referred to as infection-assortativity hereafter) indicates a tendency for infected nodes to connect with other infected nodes, and similarly for uninfected nodes (assortativ mixing by infection status). We denote the infection-assortativity of a graph with a given underlying disease distribution as $r_{\text{infection}}$.

For a graph with an underlying disease distribution generated by a stochastic SI process (see S.1), we generally expect to observe a positive $r_{\text{infection}}$, since a node can only be infected if at least one of its immediate neighbours is also infected. The exact value of $r_{\text{infection}}$ however depends on both the graph structure and the stage of outbreak progression (proportion of nodes infected) (see Fig. 3 and 4).

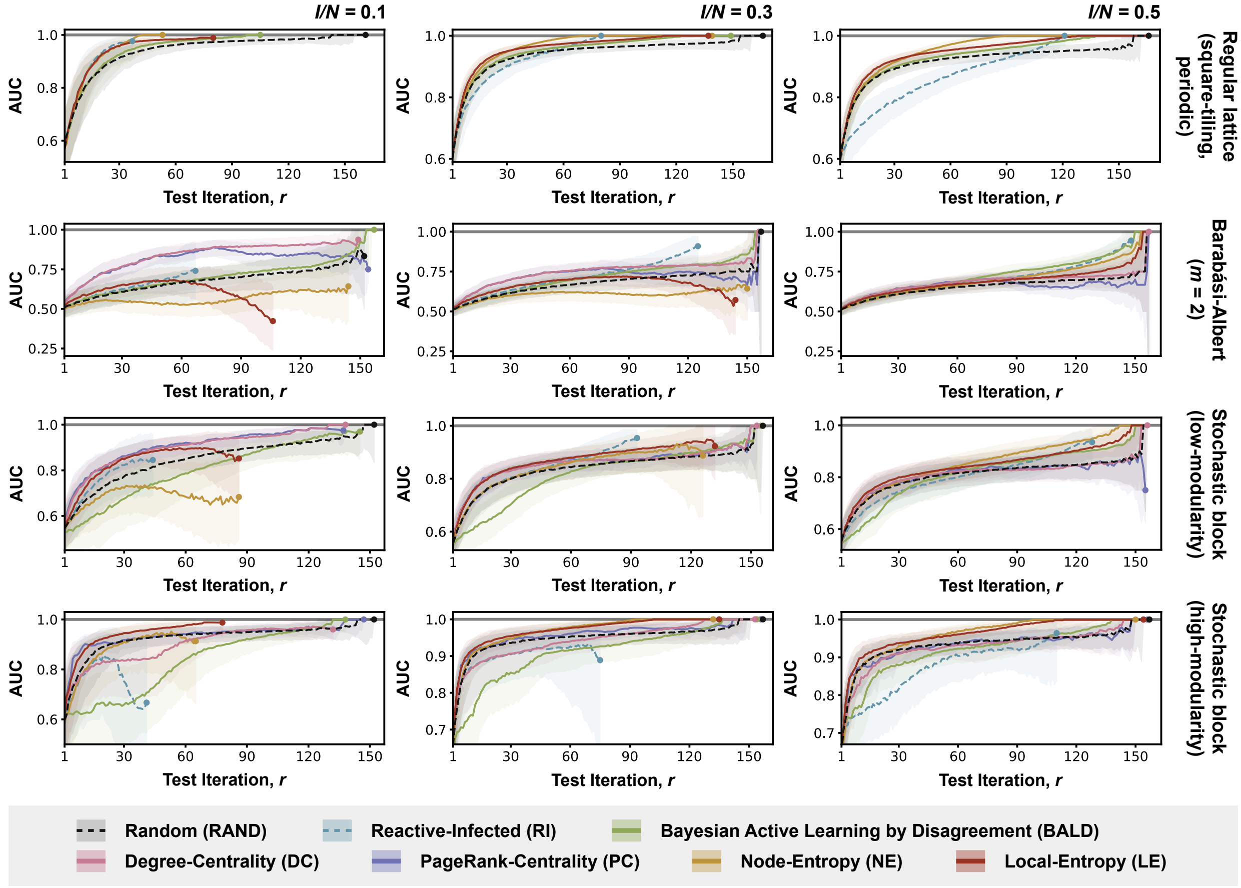


**Fig. 1.** Full results from experiments with simulated outbreaks on synthetic graphs. Each row presents results from experiments with simulated outbreaks on a different synthetic graph (as indicated by labels on the right); each column corresponds to simulated outbreaks at a different stage of outbreak progression, as measured by the proportion of nodes infected ($I/N = 0.1$, 0.3, and 0.5; as indicated by labels at the top). Each plot shows the performance of policies considered in the corresponding experiment, as measured by the AUC; the shaded region represents the interquartile range and the solid line represents the median. The performance of each policy is shown up to the median number of test iterations required for all infected nodes to be observed among agents assigned to that policy, with the AUC at this cut-off indicated by a coloured dot (unlike Fig. 3, where the performance of each policy is only shown up to the median number of test iterations required for all infected nodes to be observed among agents assigned to Reactive-Infected (RI)).

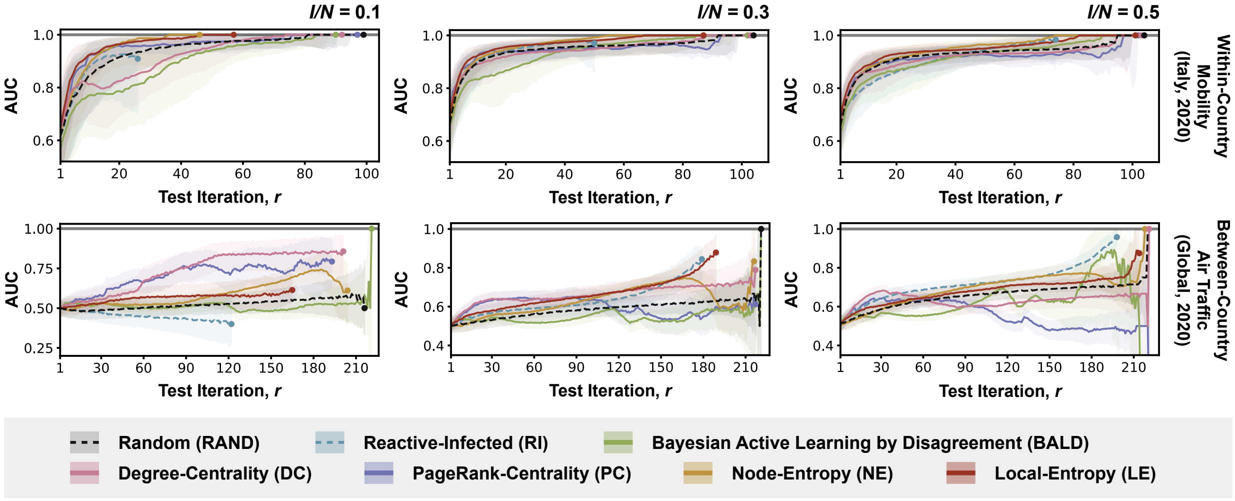


**Fig. 2.** Full results from experiments with simulated outbreaks on graphs derived from empirical human mobility data. Each row presents results from experiments with simulated outbreaks on a graph derived from a different empirical human mobility dataset (as indicated by labels on the right); each column corresponds to simulated outbreaks at a different stage of outbreak progression, as measured by the proportion of nodes infected ($I/N = 0.1$, $0.3$, and $0.5$; as indicated by labels at the top). Each plot shows the performance of policies considered in the corresponding experiment, as measured by the AUC; the shaded region represents the interquartile range and the solid line represents the median. The performance of each policy is shown up to the median number of test iterations required for all infected nodes to be observed among agents assigned to that policy, with the AUC at this cut-off indicated by a coloured dot (unlike Fig. 3, where the performance of each policy is only shown up to the median number of test iterations required for all infected nodes to be observed among agents assigned to Reactive-Infected (RI)).

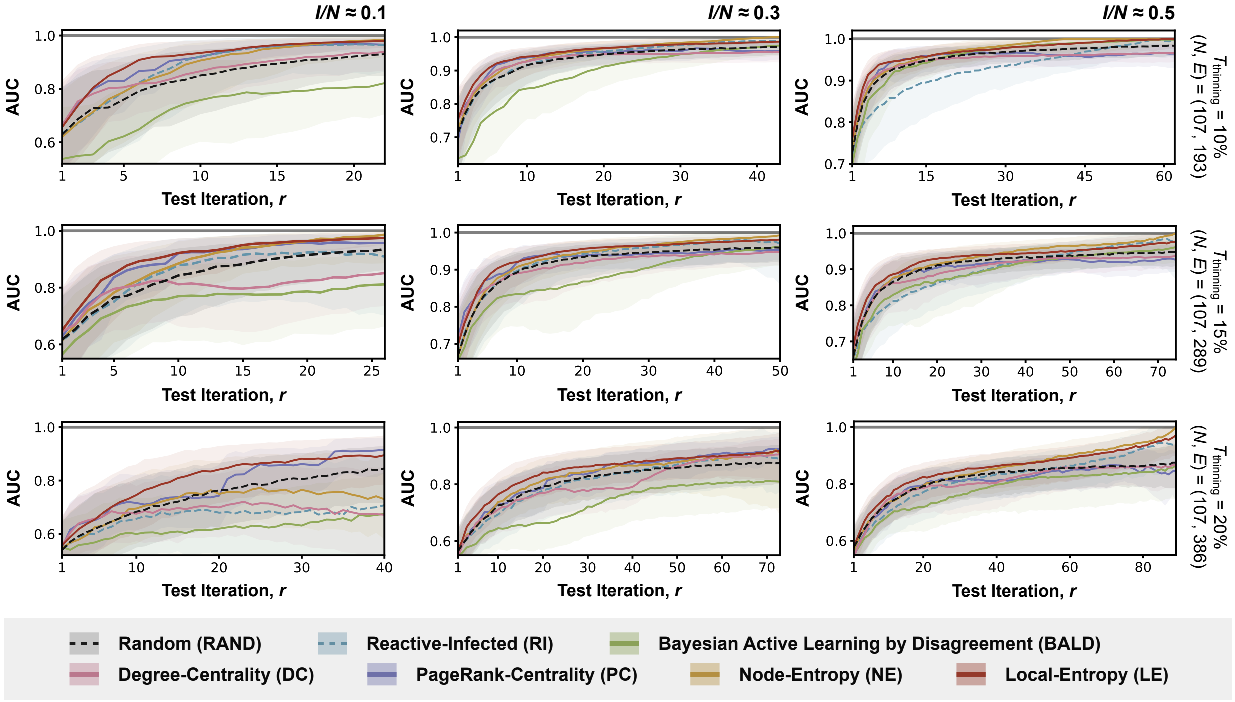


**Fig. 3.** Results from sensitivity analyses with simulated outbreaks on graphs derived from aggregated mobility data collected at provincial level in Italy. Each row corresponds to a different thinning-threshold ($T_{\text{thinning}} = 10\%$, 15%, and 20%; as indicated by labels on the right, with the number of nodes ($N$) and edges ($E$) remaining after graph-thinning also shown); each column corresponds to simulated outbreaks at a different stage of outbreak progression ($I/N = 0.1$, 0.3, and 0.5; as indicated by labels at the top). Each plot shows the performance of policies considered in the corresponding experiment, as measured by the AUC; shaded regions represent the interquartile range and the solid lines represent the median. Performance of each policy is only shown up to the median number of test iterations required for all infected nodes to be observed under the policy Reactive-Infected (RI).

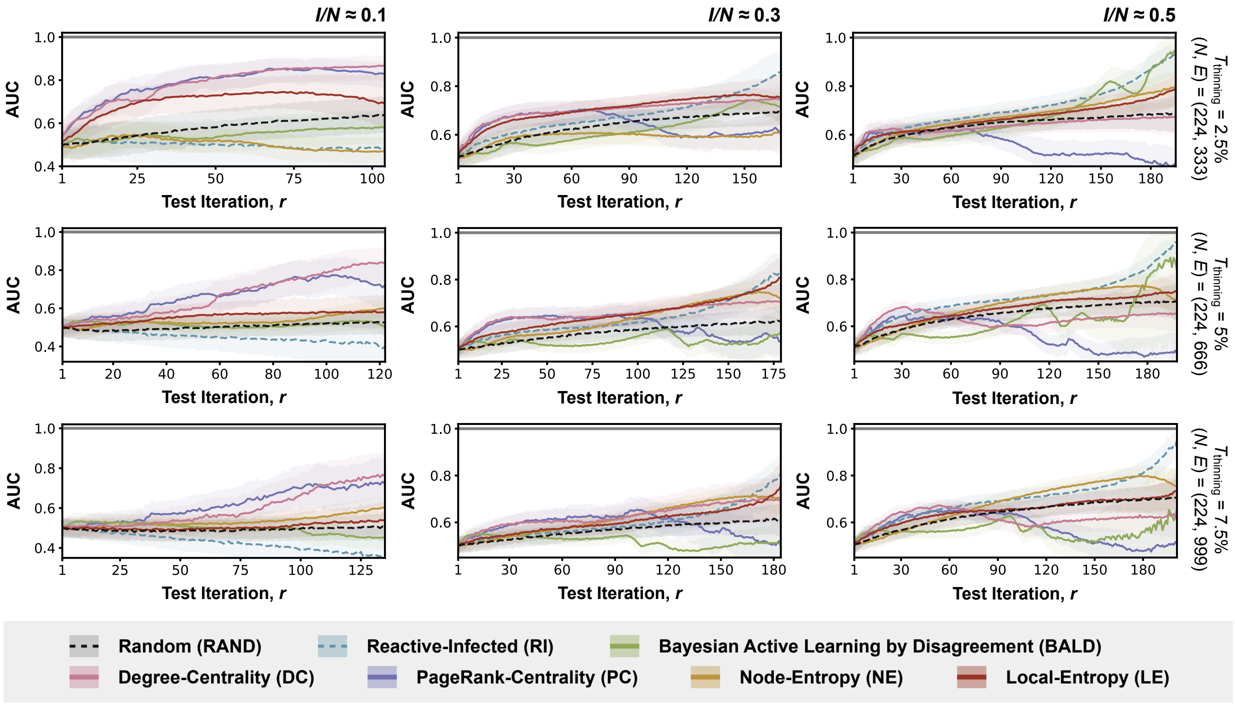


**Fig. 4.** Results from sensitivity analyses with simulated outbreaks on graphs derived from air traffic data collected at country level. Each row corresponds to a different thinning-threshold ($T_{\text{thinning}}$ = 10%, 15%, and 20%; as indicated by labels on the right, with the number of nodes ($N$) and edges ($E$) remaining after graph-thinning also shown); each column corresponds to simulated outbreaks at a different stage of outbreak progression ($I/N$ = 0.1, 0.3, and 0.5; ; as indicated by labels at the top). Each plot shows the performance of policies considered in the corresponding experiment, as measured by the AUC; shaded regions represent the interquartile range and the solid lines represent the median. Performance of each policy is only shown up to the median number of test iterations required for all infected nodes to be observed under the policy Reactive-Infected (RI).
